# DIFFERENTIAL EXPRESSION OF CB_1_ CANNABINOID RECEPTOR AND CANNABINOID RECEPTOR INTERACTING PROTEIN 1A IN LABOR

**DOI:** 10.1101/2020.12.06.20244749

**Authors:** Melissa L. Kozakiewicz, Jie Zhang, Sandra Leone-Kabler, Liliya M. Yamaleyeva, Brian C. Brost, Allyn C. Howlett

**Affiliations:** Department of Obstetrics and Gynecology, Section on Maternal-Fetal Medicine, Wake Forest School of Medicine, Medical Center Blvd., Winston-Salem, NC 27157; Department of Physiology and Pharmacology, Wake Forest School of Medicine, Medical Center Blvd., Winston-Salem, NC 27157; Department of Surgery/Hypertension and Vascular Research Center, Wake Forest School of Medicine BioTech Place, 575 N. Patterson Avenue, Suite 340, Winston-Salem, NC 27101

**Keywords:** anandamide, labor, cannabinoid receptor, endocannabinoid, myometrium, placenta

## Abstract

**Background:** The endocannabinoid system is present in multiple organ systems and is involved in smooth muscle regulation, immune function, neuroendocrine modulation and metabolism of tissues. Limited data are available regarding the presence and role of this system in reproductive tissues. Components of the endocannabinoid system have been identified in myometrial and placental tissues. However, no study has investigated differential expression of the endocannabinoid system in labor.

**Objectives:** The purpose of this study was to identify and quantify two components of the endocannabinoid system, the CB_1_ cannabinoid receptor and cannabinoid receptor interacting protein 1a, in uterine and placental tissues, and to determine if there is differential expression in tissues exposed to labor. We hypothesized that CB_1_ cannabinoid receptor concentration would be altered in uterine and placental tissue exposed to labor as compared to tissues not exposed to labor.

**Study Design:** Uterine and placental tissue samples were collected in nine laboring and 11 non-laboring women undergoing cesarean delivery. CB_1_ cannabinoid receptor and cannabinoid receptor interacting protein 1a presence and quantification were evaluated using western blot and real-time quantitative polymerase chain reaction. Statistical comparisons of laboring and non-laboring subjects were made for uterine and placental tissue using a Mann-Whitney test.

**Results:** The protein abundance of CB_1_ cannabinoid receptor in uterine tissue was significantly lower in tissues exposed to labor (p = 0.01). The protein abundance of cannabinoid receptor interacting protein 1a was lower in uterine tissue exposed to labor but did not reach statistical significance (p = 0.06). mRNA expression of CB_1_ cannabinoid receptor (p = 0.20) and cannabinoid receptor interacting protein 1a (p = 0.63) did not differ in labored compared to non-labored uterine tissues.

**Conclusions:** Our findings of diminished protein density of CB_1_ cannabinoid receptor in uterine tissue exposed to labor support the hypothesis that the endocannabinoid system plays a role in parturition. Our data add to the growing body of evidence indicating the endocannabinoid system is of importance for successful reproduction and support the need for additional research investigating this complex system as it pertains to labor.

## INTRODUCTION

Multiple pathways leading to parturition have been proposed and investigated yet a complete understanding of the signaling pathways leading to term and preterm labor has yet to be ascertained. The biology of labor is complex and includes interplay among steroid hormones, cytokines and prostaglandins affecting the maternal-fetal interface and smooth muscle regulation.^1-4^ The most accepted theory of physiologic term labor in humans is that a functional progesterone withdrawal leads to an inflammatory response, causing a cascade to the final labor pathway. It is thought that preterm labor involves several different pathologic processes that lead to a similar cascade culminating in the final labor pathway.^5^ However, decades of research targeting the inflammatory response and myometrial contractility have not resulted in effective therapy for preterm labor. Identification of a therapeutic target or marker that precedes this transition of myometrial quiescence to an active contractile state would allow innovative investigation of novel prevention or treatment strategies.

The endocannabinoid system is present in multiple organ systems and is involved in smooth muscle regulation, immune function, neuroendocrine modulation and metabolism of tissues.^6-14^ It includes CB_1_ and CB_2_ cannabinoid receptors, the endocannabinoid agonists anandamide and 2-arachidonoylglycerol (2-AG), and the enzymes that synthesize and metabolize the endocannabinoid ligands.^14-15^ The CB_1_ cannabinoid receptor is a G protein-coupled receptor with cell specific activity and is encoded by the CNR1 gene. Activation leads to coupling predominantly with G_i/o_ proteins with effects on calcium channels, mitogen-activated protein kinases and adenylyl cyclase.^14-16^ Cannabinoid receptor interacting protein 1a (CRIP1a) is a CB_1_ cannabinoid receptor-associated protein that is known to modulate CB_1_ cannabinoid receptor activity.^17,18^ The CB_1_ cannabinoid receptor specifically has been shown to influence myometrial contractility in vitro,^19^ and has been associated with the onset of labor in a mouse model.^20,21^

The ECS is of importance in sustaining the microenvironment necessary for early pregnancy success and maintenance.^22-27^ It plays a significant role in embryo development, transport and implantation as well as placentation.^22-35^ Limited data are available regarding the presence and role of this system in the mid- and late-trimesters. Components of the ECS have been identified in uterine and placental tissues.^19,35-36^ However, to our knowledge, no study has investigated differential expression of the ECS in labor. The aims of this study were to identify and quantify CB_1_ cannabinoid receptor and CRIP1a in uterine and placental tissues and to determine if there is differential expression in those tissues exposed to labor. We hypothesized that CB_1_ cannabinoid receptor concentration would be altered in uterine and placental tissue exposed to labor as compared to tissues not exposed to labor.

## MATERIALS AND METHODS

This was an observational study of women undergoing cesarean section. It was conducted after Institutional Review Board approval at Wake Forest Baptist Medical Center (Winston-Salem, NC) and Novant Forsyth Medical Center (Winston-Salem, NC). Written informed consent was obtained from each participant. Pregnant women with singleton gestations between 22 weeks, 0 days through 42 weeks, 0 days undergoing cesarean section were eligible to participate. Exclusion criteria included cannabinoid use during pregnancy, illicit drug use during pregnancy, nonsteroidal anti-inflammatory drug use within 7 days of delivery, pre-existing diabetes, pre-existing hypertension, hypertensive disorders of pregnancy, epilepsy currently treated with antiepileptic medication, intraamniotic infection and fetal anomalies.

Baseline demographics were collected from each subject’s medical record. Labor was defined as cervical change with regular contractions. Uterine and placental samples were collected at the time of cesarean section by a single surgeon. A 2 x 0.5-inch uterine sample was obtained from the superior edge of the lower uterine segment incision following delivery of the placenta. The placental samples were taken medial to the placental edge and did not include the cord insertion site. For the placental sample, the fetal surface was removed and the tissue was cut into 4 or 5 pieces (1cm sq). All tissue samples were thoroughly rinsed in sterile normal saline, placed in RNase-free sterile containers, flash frozen in the operating room and stored at −80°C immediately after delivery.

### Western Blotting

Tissue specimens (150 mg) were pulverized then homogenized with a Dremel 300 tissue homogenizer in 1.5 mL RIPA buffer (Thermo Fisher Scientific, Waltham, MA) containing proteinase inhibitor cocktail (Thermo Fisher Scientific, Waltham, MA). Samples were then centrifuged at 1,000 x g for 10 minutes at 4□C. The supernatant was centrifuged at 25,000 x g for 20 min at 4□C. Protein concentrations were measured using a BCA assay.^37^ Samples were prepared in the loading buffer at 2 µg/µL and heated at 65□C for 8 min. Samples (60 µg) were loaded onto a 4-20% Novex wedgewell gel (Thermo Fisher Scientific, Waltham, MA) and electrophoresed at 60 v for 5 min followed by 100 v for 90 min, and transferred to a polyvinylidene fluoride membrane (Millipore, Burlington, MA). Blots were allowed to dry completely and were stored in the dark at room temperature until antibody incubation. Blots were rewetted in methanol for 15 sec, rinsed for 1 min in PBS, and blocked for 60 min with Odyssey blocking buffer (LI-COR Biosciences Lincoln, NE) at 21-23°C. Blots were probed with primary antibodies (4°C for 18 hours) for CB_1_ cannabinoid receptor (Catalog # EB10961; Everest Biotech, Oxfordshire, UK), CRIP1a (Catalog # sc-515504; Santa Cruz Biotechnology, Santa Cruz, CA) and β-Actin (Catalog # 66009; Proteintech, Rosemont, IL). Blots were washed and incubated with an appropriate Li-COR IR dye-conjugated secondary antibody. Bands were imaged using the Odyssey CLx Infrared Imaging System and quantified by densitometry using Image Studio software (LI-COR Biosciences, Lincoln, NE). Band densities were normalized to β-actin as a loading control and then normalized to the mean of all samples being assigned a value of 1.0.

### Quantitative Real-Time Polymerase Chain Reaction (qPCR)

Ribonucleic acid (RNA) was isolated using the RNeasy mini kit (Qiagen, Santa Clarita, CA). RNA yield and purity were determined spectrophometrically using a NanoDrop (Thermo Fisher Scientific, Waltham, MA). The RNA was reverse transcribed to cDNA using a high capacity cDNA kit (Catalog # 4368814, Thermo Fischer Scientific, Waltham, MA).

GAPDH and HPRT1 were first evaluated for variability between sample groups. Because their expression did not differ between labored and non-labored samples in uterine or placental tissues, these genes were used for normalization in the present studies. The primers used were as follows: *CNR1* 5’-AGCAGACCAGGTGAACATTAC-3’ and 3’-GACCATGAAACACTCTATG-5’; *CRIP1a* 5’- CCAGTTGTTCTCGGTCATACTT-3’ and 3’-AAAGAGCGGAGCTGTTTATAGG-5’; *GAPDH* 5’-ACATCGCTCAGACACCATG-3’ and 3’-TGTAGTTGAGGTCAATGAAGGG- 5’; *HPRT1* 5’-TTGTTGTAGGATATGCCCTTGA-3’ and 3’- GCGATGTCAATAGGACTCCAG-5’ (Integrated DNA Technologies, Coralville, IA). cDNA was diluted to 5 ng/mL. qPCR was performed using SYBR Green Master Mix (Catalog # QP001-01; GeneCopoeia, Rockville, MD) in triplicate with the Applied Biosystems StepOne Real-Time PCR System (Applied Biosystems, Foster City, CA). The comparative threshold cycle method was used to calculate relative gene expression.^38^

### Statistical Analysis

Statistical comparisons for uterine and placental tissue were made using a Mann-Whitney test, looking at differences in receptor concentration among tissues exposed to labor compared to those not exposed to labor. Significant difference was considered to be p<0.05. All analyses were performed using GraphPad Prism version 8.3.0 for Windows (GraphPad Software, San Diego, CA, USA). Data are presented as mean ±SEM. Sample size calculation was not performed as there was no published literature to determine what effect size would be significant.

## RESULTS

Twenty healthy subjects were enrolled in the study. The median gestational age was 38 weeks, 5 days in the labor group (n = 9) and 39 weeks, 0 days in the non-labor group (n = 11). Additional characteristics are listed in Table 1. There was no significant difference in age, body mass index, race, gestational age, tobacco use or birth weight. There was a significant difference in parity with the labor group having more nulliparous subjects (p = 0.02). Cesarean deliveries were performed in the non-labor group for history of cesarean section with desire for repeat cesarean, previous shoulder dystocia, breech presentation and fetal growth restriction with non-reassuring antenatal testing. Cesarean deliveries were performed in the labor group for non-reassuring fetal heart rate tracing, history of cesarean section with desire for repeat, and arrest of descent. All subjects received either epidural or spinal anesthesia.

### Uterus

CB_1_ cannabinoid receptor and CRIP1a were both identified in uterine samples. The protein abundance of CB_1_ cannabinoid receptor in uterine tissues was significantly lower in laboring subjects compared to non-laboring subjects (Figure 1). Band intensity of CB_1_ cannabinoid receptor normalized to β-actin was 0.79 ± 0.118 (n = 9) for all laboring subjects and 1.19 ± 0.093 (n = 11) for all non-laboring subjects (p = 0.01). Relative band intensity of CRIP1a was 0.76 ± 0.125 (n = 9) for all laboring subjects and 1.17 ± 0.126 (n = 11) for all non-laboring subjects (p *=* 0.06). Subgroup analysis of term laboring subjects (n = 6) compared to term non-laboring subjects (n = 9) identified less CB_1_ cannabinoid receptor protein in tissue exposed to labor compared to tissue not exposed to labor (p = 0.02). CRIP1a protein abundance was also significantly lower in labored uterine tissue in the term subgroup (p = 0.03). While there were significantly more nulliparous subjects in the laboring group, there was no significant difference in the CB_1_ cannabinoid receptor (p = 0.17) or CRIP1a (p = 0.38) protein density between nulliparous and multiparous laboring subjects in uterine tissue.

**Figure 1:**
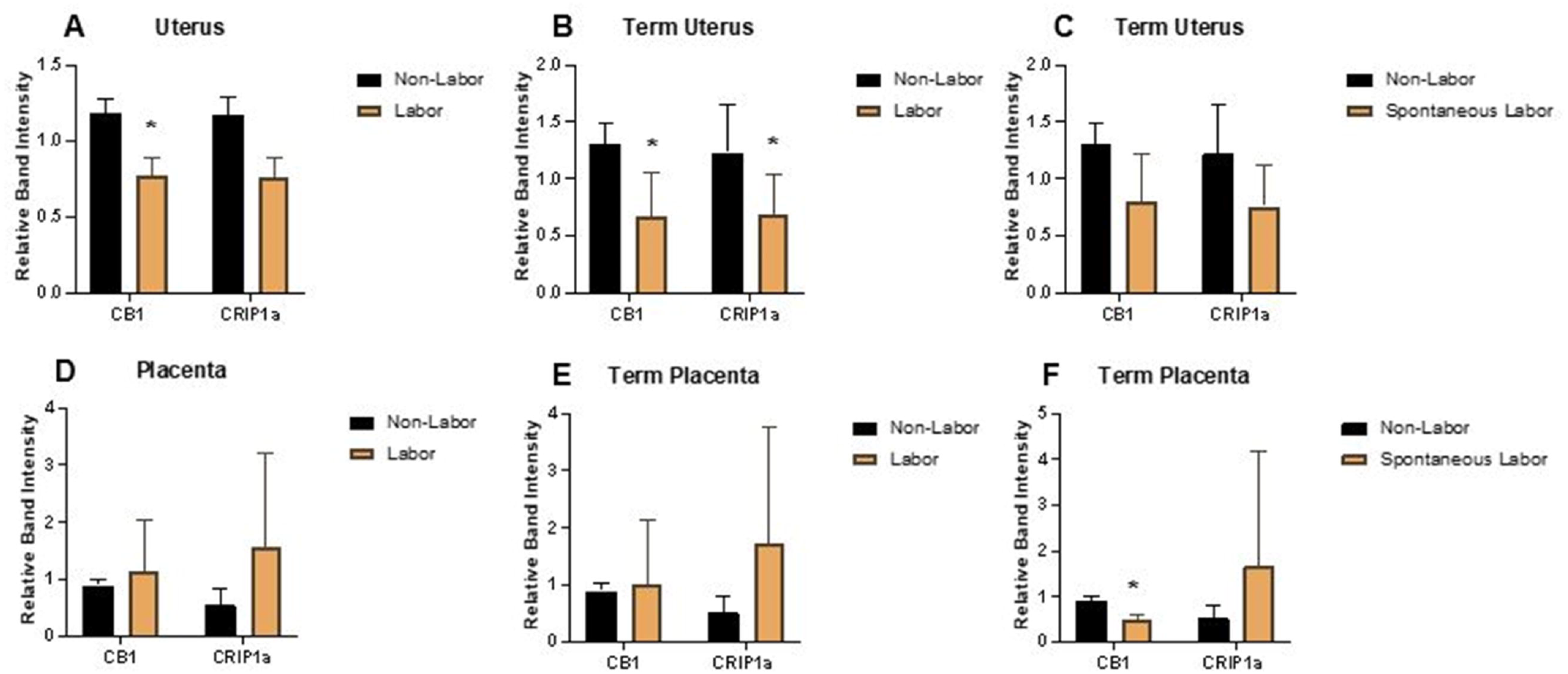
Western blot band intensities of CB_1_ cannabinoid receptor and CRIP in laboring and non-laboring uterus and placenta. ***A, Uterus*** *-* Relative band intensity of CB_1_ receptor (*p =* 0.01) and CRIP1a (*p =* 0.06) for all subjects in the laboring (n = 9) and non-laboring (n = 11) groups. ***B, Uterus*** *-* Relative band intensity of CB_1_ receptor (*p* 0.02) and CRIP1a (*p* = 0.03) for term laboring (n = 6) and non-laboring (n = 9) subjects. ***C, Uterus*** *-* Relative band intensity of CB_1_ receptor (*p* = 0.11) and CRIP1a (*p* = 0.11) for term spontaneously laboring (n = 4) and non-laboring (n = 9) subjects. ***D, Placenta*** *-* Relative band intensity of CB_1_ receptor (*p =* 0.87) and CRIP1a (*p =* 0.05) for all subjects in the laboring (n = 9) and non-laboring (n = 11) groups. ***E, Placenta*** *-* Relative band intensity of CB_1_ receptor (*p* = 0.14) and CRIP1a (*p* = 0.22) for term laboring (n = 6) and non-laboring (n = 9) subjects. ***F, Placenta*** *-* Relative band intensity of CB_1_ receptor (*p* < 0.01) and CRIP1a (*p* = 0.71) for term spontaneously laboring (n = 4) and non-laboring (n = 9) subjects. Data are presented as means ±SEM, and statistical comparisons were made using a Mann-Whitney test, with significant difference (*) considered to be p<0.05.

**Figure 2:**
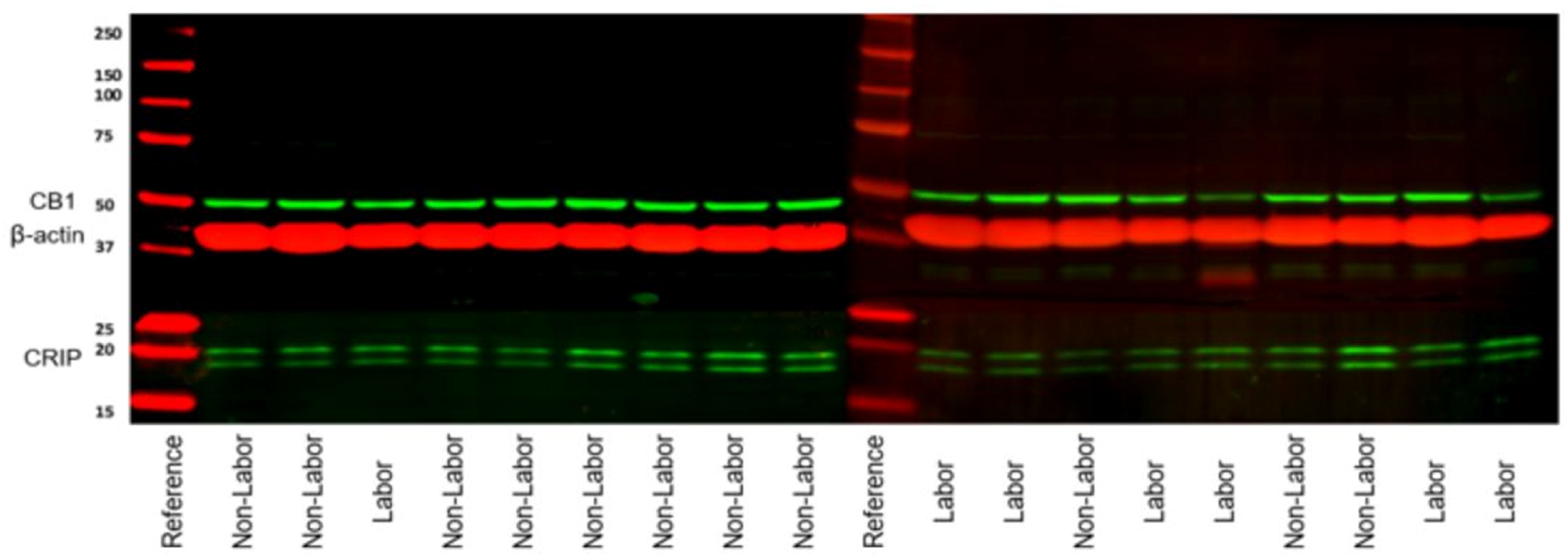
Western Blot Images. CB_1_ and CRIP1a immunoblots were visualized using the LI-COR Odyssey system. Bands were normalized to β-actin loading control.

There was no significant difference in mRNA expression of CB_1_ cannabinoid receptor (p = 0.20) or CRIP1a (p = 0.63) in labored compared to non-labored uterine tissues (Figure 3). Subgroup analyses including only term subjects and term subjects in spontaneous labor also showed no significant difference in mRNA expression of CB_1_ cannabinoid receptor or CRIP1a.

**Figure 3:**
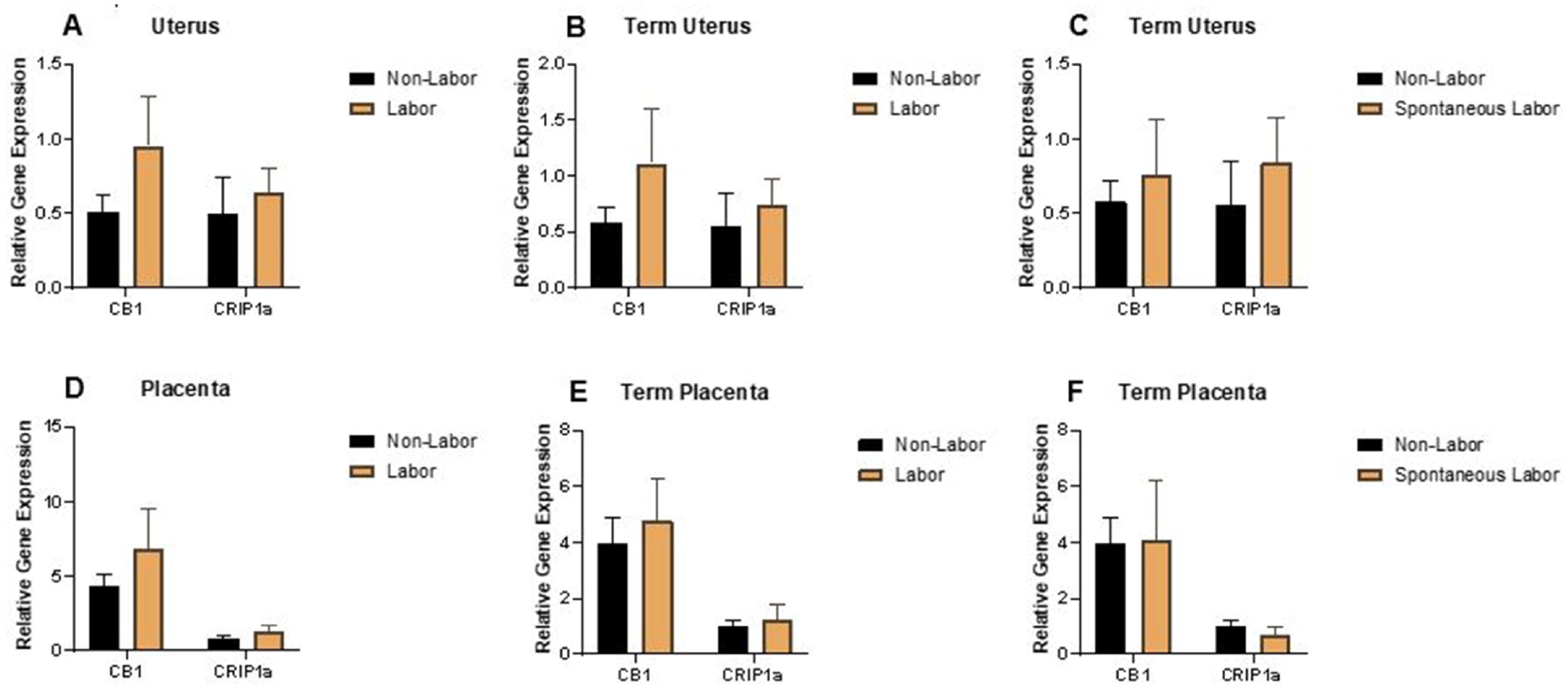
mRNA expression of CB_1_ cannabinoid receptor and CRIP1a using qPCR did not differ in uterine or placental tissues exposed to labor. ***A, Uterine*** *–* mRNA expression of CB_1_ receptor (*p =* 0.37) and CRIP1a (*p =* 0.08) for all subjects in the laboring (n = 9) and non-laboring (n = 11) groups. ***B, Uterine*** *–* mRNA expression of CB_1_ receptor (*p* = 0.61) and CRIP1a (*p* = 0.22) for term laboring (n = 6) and non-laboring (n = 9) subjects. ***C, Uterine*** *–* mRNA expression of CB_1_ receptor (*p* = 0.94) and CRIP1a (*p* = 0.08) for term spontaneously laboring (n = 4) and non-laboring (n = 9) subjects. ***D, Placenta*** *–* mRNA expression of CB_1_ receptor (*p =* 0.88) and CRIP1a (*p =* 0.66) for all subjects in the laboring (n = 9) and non-laboring (n = 11) groups. ***E, Placenta*** *–* mRNA expression of CB_1_ receptor (*p* = 0.78) and CRIP1a (*p* = 0.86) for term laboring (n = 6) and non-laboring (n = 9) subjects. ***F, Placenta*** *–* mRNA expression of CB_1_ receptor (*p* = 0.83) and CRIP1a (*p* = 0.26) for term spontaneously laboring (n = 4) and non-laboring (n = 9) subjects. The comparative threshold cycle method was used to calculate relative gene expression. Gene expression was normalized to a pooled reference sample. Data are presented as fold difference mean ±SEM, and statistical comparisons were made using a Mann-Whitney test, with significant difference (*) considered to be p<0.05.

### Placenta

CB_1_ cannabinoid receptor and CRIP1a were both identified in human placental samples. There was no significant difference in band density of the western blots when comparing the labor and non-labor groups (Figure 1). Band intensity of CB_1_ cannabinoid receptor normalized to β-actin was 1.13 ± 0.306 (n = 9) for all laboring patients and 0.89 ± 0.035 (n = 11) for all non-laboring patients (p = 0.40). Relative band intensity of CRIP1a was 1.56 ± 0.549 (n = 9) for all laboring patients and 0.55 ± 0.083 (n = 11) for all non-laboring patients (p *=* 0.06). No significant difference in CB_1_ cannabinoid receptor (p = 0.36) or CRIP1a (p = 0.30) mRNA expression was identified between the two groups (Figure 3).

## DISCUSSION

Our data confirm that the components of the ECS are present in human uterine and placental tissues by using both western blot to confirm protein presence and qPCR to confirm mRNA presence. The most prominent finding was that CB_1_ cannabinoid receptor levels are significantly diminished in uterine tissue exposed to labor. However, no significant difference was identified between labored and non-labored placental tissue. These findings add to the evidence that the ECS is involved in uterine and placental function.

Previous studies have demonstrated cannabinoid receptor presence in myometrial and placental tissues.^19, 35-36^ A study evaluating myometrial cells obtained from segments of term human myometrial tissue identified CB_1_ and CB_2_ cannabinoid receptors. ^19^ In the same study, the effects of cannabinoids on uterine contractility were evaluated in vitro and a CB_1_ cannabinoid receptor-mediated relaxant effect of endogenous and exogenous cannabinoids on myometrial contractility was demonstrated.^19^ Additionally, a mouse model has previously been used to evaluate the effect of CB_1_ cannabinoid receptor inactivation on parturition.^20^ Mice lacking the CB_1_ cannabinoid receptor were found to experience earlier onset of labor compared to wild type mice.^20^ In wild type mice in the same study, CB_1_ cannabinoid receptor silencing in late gestation also resulted in labor.^20^ Our findings of reduced CB_1_ cannabinoid receptor protein in laboring uterine tissue are concordant with these previously published findings.

Despite decades of research, preterm labor remains a significant cause of maternal-fetal morbidity and economic strain.^39-40^ Complications related to premature birth include neurodevelopmental disorders, retinopathy of prematurity, bronchopulmonary dysplasia and hearing impairment among others. Physiologic and pathologic mechanisms leading to the initiation of the final labor pathway have been implicated in the etiology of term and preterm birth, but a clear understanding of the pathophysiology has not been ascertained.^1-5,39-44^ The major challenge in the development of treatment and preventative therapies is the lack of a clear understanding of the processes responsible for shifting the myometrium from quiescence to an active contractile state. Identifying the ECS as a system that contributes to events preceding the labor cascade may open the door to therapeutic modalities not previously studied and would help to better understand the physiology of labor.

Current treatment of preterm labor targets stimulation of fetal maturity with corticosteroids, magnesium sulfate for fetal neuroprotection and the consideration of tocolytic medications targeting uterine contractility.^39^ Various classes of medications to decrease uterine contractions have been studied including calcium channel blockers, selective beta 2-adregergic agonists, prostaglandin-synthase or cyclooxygenase (COX) inhibitors and oxytocin receptor antagonists.^41-44^ Despite their frequent use, no tocolytic therapy has been shown to independently improve neonatal outcomes. First-line tocolytic medications currently recommended include nifedipine (calcium channel blocker) and indomethacin (non-selective COX inhibitor).^39^ Their short-term use is indicated to allow for the administration of corticosteroids, magnesium sulfate for fetal neuroprotection and for transfer to a tertiary care center with neonatology capabilities. Indomethacin has been studied as a tocolytic therapy since the 1970’s after it was noted to inhibit prostaglandin activity.^44^ Interestingly, a recent study has identified indomethacin as being a positive allosteric modulator of the CB_1_ cannabinoid receptor.^45^ Modulation of CB_1_ cannabinoid receptor signaling pathways has been a target of investigation with significant overlap of the prostaglandin and endocannabinoid pathways noted.^45-46^ Our data demonstrating lower levels of CB_1_ cannabinoid receptor in labored uterine tissue suggest that downregulation of the ECS during labor could be important for the mechanisms allowing labor to proceed. Allosteric modulators of the CB_1_ cannabinoid receptor are being investigated as possible therapy for various pathologic disorders in non-reproductive organ systems.^46-48^ Further investigation of ECS-related pathways may elucidate targets for clinical research pertaining to parturition.

Our findings have significant research implications; specifically, they warrant further investigation of the ECS as a system intimately involved in the labor process. One must contemplate if this system contributes to the maintenance of uterine quiescence. While we have determined there is a significant decrease in CB_1_ cannabinoid receptor in laboring uterine tissue, there are other components of the ECS still to be studied in this population. The endogenous lipid ligands anandamide and 2-AG bind to the CB_1_ and CB_2_ cannabinoid receptors.^14^ Plasma anandamide levels have been correlated with risk of spontaneous pregnancy loss in the first trimester.^30-32^ Both anandamide and 2-AG have been identified as being of critical importance in early pregnancy events.^22-32^ Furthermore, plasma levels of anandamide have been shown to increase at term and dramatically increase in labor.^49^ Recent research has included the endocannabinoids, their precursors and their pathways of degradation as targets for prediction of preterm labor.^50^ Anandamide is primarily made from *N-*arachidonyl phosphatidylethanolamine (NAPE) by NAPE-hydrolyzing phospholipase D although other pathways can also contribute to its synthesis.^9,10^ It is degraded by the enzyme fatty acid amide hydrolase (FAAH). The activity of FAAH has been shown to be an important factor in early pregnancy success.^32,35^ It has been shown that low levels of anandamide correlate with high levels of FAAH at uterine implantation sites suggesting this is an important factor for successful pregnancy implantation.^22^ FAAH protein expression as it relates to CB_1_ cannabinoid receptor expression has been described using immunohistochemistry in term membranes and placentas.^36^ Furthermore, mice without FAAH and thus with increased anandamide have been shown to be more susceptible to inflammation-induced preterm labor regardless of progesterone levels.^21^

In our study, while the protein levels of CB_1_ cannabinoid receptor differed in the uterine tissue exposed to labor compared to that not exposed to labor, mRNA expression did not vary by qPCR. Investigation of the mechanisms explaining the variation in CB_1_ cannabinoid receptor and CRIP1a protein abundance without a change in mRNA expression is warranted. Mechanisms of interest include receptor desensitization and protein degradation.

To our knowledge, the current study is the first to quantify the CB_1_ cannabinoid receptor and CRIP1a in laboring uterine and placental tissues. It included a fairly homogenous population of women without significant co-morbidities, and all samples were collected consistently by a single surgeon. A limitation of this study is that the laboring group did include a small number of patients being induced or augmented with oxytocin. However, the vast majority were spontaneously laboring patients. Also, the underlying factors necessitating cesarean section as the mode of delivery could have impacted our results.

## Conclusions

There exists significant overlap between prostaglandins, cytokines, steroid hormones and the ECS. A growing body of evidence indicates that appropriate interaction between these pathways is important for successful reproduction and normal labor. Our data underscore the need for additional research investigating this complex system as it pertains to labor.

## Data Availability

The data that support the findings of this study are available from the corresponding author, [MLK], upon reasonable request.

## Acknowledgements

The authors wish to acknowledge Christina Tulbert and Dr. Jorge Figueroa for their assistance in acquisition of equipment for the study.

## REFERENCES

1. Olson DM. The role of prostaglandins in the initiation of parturition. Best Pract Res Clin Obstet Gynaecol 2003;17:717–30.

2. Shynlova O, Tsui P, Jaffer S, et al. Integration of endocrine and mechanical signals in the regulation of myometrial functions during pregnancy and labour. Eur J Obstet Gynecol Reprod Biol 2009;144 Suppl 1:S2–10.

3. Renthal NE, Williams KC, Montalbano AP, et al. Molecular regulation of parturition: a myometrial perspective. Cold Spring Harb Perspect Med 2015;5:a023069.

4. Lee Y, Sooranna SR, Terzidou V, et al. Interactions between inflammatory signals and the progesterone receptor in regulating gene expression in pregnant human uterine myocytes. J Cell Mol Med 2012;16:2487–503.

5. Romero R, Dey SK, Fisher SJ. Preterm labor: one syndrome, many causes. Science 2014;345:760–5.

6. De Laurentiis A, Fernandez-Solari J, Mohn C, et al. The hypothalamic endocannabinoid system participates in the secretion of oxytocin and tumor necrosis factor-alpha induced by lipopolysaccharide. J Neuroimmunol 2010;221(1-2):32–41.

7. Mallat A, Teixeira-Clerc F, Lotersztajn S. Cannabinoid signaling and liver therapeutics. J Hepatol 2013;59:891–6.

8. Hiley CR. Endocannabinoids and the heart. J Cardiovasc Pharmacol 2009;53:267–76.

9. Pulgar VM, Yamaleyeva LM, Varagic J, et al. Increased angiotensin II contraction of the uterine artery at early gestation in a transgenic model of hypertensive pregnancy is reduced by inhibition of endocannabinoid hydrolysis. Hypertension 2014;64:619–25.

10. Cota D. The role of the endocannabinoid system in the regulation of hypothalamic-pituitary-adrenal axis activity. J Neuroendocrinol 2008;20 Suppl 1:35–8.

11. Maia J, Almada M, Silva A, et al. The endocannabinoid system expression in the female reproductive tract is modulated by estrogen. J Steroid Biochem Mol Biol 2017;174:40–7.

12. Leishman E, Cornett B, Spork K, et al. Broad impact of deleting endogenous cannabinoid hydrolyzing enzymes and the CB1 cannabinoid receptor on the endogenous cannabinoid-related lipidome in eight regions of the mouse brain. Pharmacol Res 2016;110:159–72.

13. Leishman E, Mackie K, Luquet S, et al. Lipidomics profile of a NAPE-PLD KO mouse provides evidence of a broader role of this enzyme in lipid metabolism in the brain. Biochim Biophys Acta 2016;1861:491–500.

14. Howlett AC, Blume LC, Dalton GD. CB1 cannabinoid receptors and their associated proteins. Curr Med Chem 2010;17:1382–93.

15. Howlett AC. The cannabinoid receptors. Prostaglandins Other Lipid Mediat 2002;68-69:619–31.

16. Brighton PJ, Marczylo TH, Rana S, et al. Characterization of the endocannabinoid system, CB1 receptor signaling and desensitization in human myometrium. Br J Pharmacol 2011;164:1479–94.

17. Smith TH, Blume LC, Straiker A, et al. Cannabinoid receptor-interacting protein 1a modulates CB1 receptor signaling and regulation. Mol Pharmacol 2015;87:747–65.

18. Booth WT, Walker NB, Lowther WT, et al. Cannabinoid receptor interacting protein 1a (CRIP1a): function and structure. Molecules 2019;24:3672. doi: 10.3390/molecules24203672.

19. Dennedy MC, Friel AM, Houlihan DD, et al. Cannabinoids and the human uterus during pregnancy. Am J Obstet Gynecol 2004;190:2–9.

20. Wang H, Xie H, Dey SK. Loss of cannabinoid receptor CB1 induces preterm birth. PLoS One 2008;3:e3320.

21. Sun X, Deng W, Li Y, Tang S, et al. Sustained endocannabinoid signaling compromises decidual function and promotes inflammation-induced preterm birth. J Biol Chem 2016;291:8231–40.

22. Wang H, Xie H, Sun X, et al. Differential regulation of endocannabinoid synthesis and degradation in the uterus during embryo implantation. Prostaglandins Other Lipid Mediat 2007;83:62–74.

23. Schmid PC, Paria BC, Krebsbach RJ, et al. Changes in anandamide levels in mouse uterus are associated with uterine receptivity for embryo implantation. Proc Natl Acad Sci U S A 1997;93:188–92.

24. Das SK, Paria BC, Chakraborty I, et al. Cannabinoid ligand-receptor signaling in the mouse uterus. Proc Natl Acad Sci U S A 1995;92:4332–6.

25. Paria BC, Wang SX, Schmid PC, et al. Dysregulated cannabinoid signaling disrupts uterine receptivity for embryo implantation. J Biol Chem 2001;276:20523–8.

26. Paria BC, Wang H, Dey SK. Endocannabinoid signaling in synchronizing embryo development and uterine receptivity for implantation. Chem Phys Lipids 2002;121:201–10.

27. Paria BC, Dey SK. Ligand-receptor signaling with endocannabinoids in preimplantation embryo development and implantation. Chem Phys Lipids 2000;108:211–20.

28. Sun X, Dey SK. Endocannabinoid signaling in female reproduction. ACS Chem Neurosci 2012;3(5):349–55

29. Walker OS, Holloway AC, Raha S. The role of the endocannabinoid system in female reproductive tissues. J Ovarian Res 2019;12:3. doi: 10.1186/s13048-018-0478-9.

30. Habayeb OM, Taylor AH, Finney M, et al. Plasma anandamide concentration and pregnancy outcome in women with threatened miscarriage. JAMA 2008;299:1135–6.

31. Maccarrone M, Bisogno T, Valensise H, et al. Low fatty acid amide hydrolase and high anandamide levels are associated with failure to achieve an ongoing pregnancy after IVF and embryo transfer. Mol Hum Reprod 2002;8:188–95.

32. Maccarrone M, Valensise H, Bari M, et al. Relation between decreased anandamide hydrolase concentrations in human lymphocytes and miscarriage. Lancet 2000;355:1326–9.

33. Meccariello R, Battista N, Bradshaw HB, et al. Updates in reproduction coming from the endocannabinoid system. Int J Endocrinol 2014;2014:412354. doi: 10.1155/2014/412354.

34. Cecconi S, Rapino C, Di Nisio V, et al. The (endo)cannabinoid signaling in female reproduction: what are the latest advances? Prog Lipid Res 2020 Jan;77:101019.

35. Trabucco E, Acone G, Marenna A, et al. Endocannabinoid system in first trimester placenta: low FAAH and high CB1 expression characterize spontaneous miscarriage. Placenta 2009;30:516–22.

36. Park B, Gibbons HM, Mitchell MD, et al. Identification of the CB1 cannabinoid receptor and fatty acid amide hydrolase (FAAH) in the human placenta. Placenta 2003;24:473–8.

37. Smith PK, Krohn RI, Hermanson GT, et al. Measurement of protein using bicinchoninic acid. Anal Biochemo 1985;150:76–85.

38. Livak KJ, Schmittgen TD. Analysis of relative gene expression data using real-time quantitative PCR and the 2(-Delta Delta C(T)) method. Methods 2001;25: 402–8.

39. American College of Obstetricians and Gynecologists’ Committee on Practice Bulletins – Obstetrics. Practice bulletin no. 171: management of preterm labor. Obstet Gynecol 2016;128:e155–64.

40. Goldenberg RL, Culhane JF, Iams JD, et al. Epidemiology and causes of preterm birth. Lancet 2008;371:75–84.

41. Navahe R, Berghella V. Tocolysis for acute preterm labor: where have we been, where are we now and where are we going? Amer J Perinatol 2016;33:229–35.

42. Younger JD, Reitman E, Gallos G. Tocolysis: present and future treatment options. Semin Perinatol 2017;41:493–504.

43. Reinebrant HE, Pileggi-Castro C, Romero CLT, et al. Cyclo-oxygenase (COX) inhibitors for treating preterm labour. Cochrane Database of Systematic Reviews 2015, Issue 6. Art. No.: CD001992. DOI: 10.1002/14651858.CD001992.pub3.

44. Zuckerman H, Reiss U, Rubinstein I. Inhibition of human premature labor by indomethacin. Obstet Gynecol 1974;44:787–92.

45. Laprairie RB, Mohamed KA, Zagzoog A, et al. Indomethacin enhances type 1 cannabinoid receptor signaling. Front Mol Neurosci 2019;12:257. doi: 10.3389/fnmol.2019.00257.

46. Nguyen T, Li JX, Thomas BF, Wiley JL, et al. Allosteric modulation: An alternate approach targeting the cannabinoid CB1 receptor. Med Res Rev 2017;37:441–74.

47. Laprairie RB, Bagher AM, Kelly ME, et al. Cannabidiol is a negative allosteric modulator of the cannabinoid CB1 receptor. Br J Pharmacol 2015;172:4790–805.

48. Ignatowska-Jankowska BM, Baillie GL, Kinsey S, et al. A cannabinoid CB1 receptor-positive allosteric modulator reduces neuropathic pain in the mouse with no psychoactive effects. Neuropsychopharmacology 2015;40:2948–59.

49. Habayeb OM, Taylor AH, Evans MD, et al. Plasma levels of the endocannabinoid anandamide in women—a potential role in pregnancy maintenance and labor? J Clin Endocrinol Metab 2004;89:5482–7.

50. Bachkangi P, Taylor AH, Bari M, et al. Prediction of preterm labour from a single blood test: The role of the endocannabinoid system in predicting preterm birth in high-risk women. Eur J Obstet Gynecol Reprod Biol 2019;243:1–6.

51. Accialini P, Etcheverry T, Malbrán MN, et al. Anandamide regulates oxytocin/oxytocin receptor system in human placenta at term. Placenta 2020;93:23–5.

52. Baumbach J, Shi SQ, Shi L, et al. Inhibition of uterine contractility with various tocolytics with and without progesterone: in vitro studies. Am J Obstet Gynecol 2012;206:254.e1-5.

53. Al-Zoubi R, Morales P, Reggio PH. Structural insights into CB1 receptor biased signaling. Int J Mol Sci 2019;20:1837. doi: 10.3390/ijms20081837.

